# Evidence for Mother-to-Child Transmission of HIV-2 in Uganda: A Retrospective Analysis

**DOI:** 10.1101/2024.01.10.23299160

**Authors:** Grace Esther Kushemererwa, Acellam Sam, Oreen Kemigisha, Linda Kisakye Nabitaka, Christine Namulindwa, Cordelia Katureebe, Susan Nabadda, Isaac Ssewanyana

**Affiliations:** Central Public Health Laboratories, Ministry of Health Uganda; Ministry of Health Uganda

## Abstract

**Background:** HIV-2 prevalence is poorly understood outside its West African epicenter, contributing to gaps in global epidemiolocal understanding. Cases have been identified in countries like India, Europe, and the Americas, largely due to migration and travel. With a clinical presentation marked by lower viremia and reduced transmission risk, HIV-2 progresses to AIDS more gradually than HIV-1. However, it can still lead to significant health issues. The overlap in diagnostic profiles for HIV-1 and HIV-2 often results in the under-recognition of the latter in areas where HIV-1 is dominant. This under-detection poses challenges in eradicating the epidemic, as current testing protocols may not adequately differentiate between the two strains. In Uganda, we recently transitioned to the Roche Cobas 8800/6800 using the Cobas^®^ HIV-1/HIV-2 Qualitative nucleic acid test, for the early infant diagnosis of HIV. The platform has the ability to differentiate between HIV-1 and HIV-2 to detect presence of HIV-2 in DBS, Serum, Plasma and whole blood samples. This represents a pivotal shift toward refining early infant diagnosis and underscores the need for nuanced surveillance to address the distinct epidemiology of HIV-2.

**Method:** Twenty-four thousand six hundred and nineteen (24,619) Dry blood spots (DBS) collected from infants under 18 months old, all born to mothers living with HIV AIDs in Uganda were tested according to the Uganda consolidated guidelines for HIV prevention and treatment. These were tested for routine early infant diagnosis (EID) at the centralized reference lab as per the Uganda national HIV consolidated guideline. Testing was performed using the Cobas^®^ HIV-1/HIV-2 Qualitative nucleic acid test for use on the Cobas^®^ 5800/6800/8800 Systems. This advanced testing method not only detects HIV but also differentiates between HIV-1 and HIV-2, which is crucial for accurate diagnosis and treatment. It’s a reliable test, with a specificity confirmed to be 100% (95% confidence limit: ≥ 99.5%) and it is both FDA approved and WHO Pre-qualified.

**Results:** Of the 24,619 DBS tested between May and November 2023, 466 were confirmed positive (1.9%). Four (4) of the 466 (0.9%) were confirmed HIV-2. These 4 are from Kampala (1), Kagadi (1), Namutumba (1), and Oyam (1) districts. The HIV-2 positive samples had higher CT values (39.2, 40.33, 40.35, 44.62) compared to the average less than 30 for the HIV-1 positive samples and are representative of the 95^th^ percentile. The significance of the CT (cycle threshold) values obtained for HIV-2 positive samples is crucial for understanding the viral load and transmission risk. Compared to HIV-1, HIV-2 is generally associated with lower viral loads, as indicated by higher CT values in PCR testing. This lower viral load is a key factor in the reduced transmissibility of HIV-2 compared to HIV-1. In the context of mother-to-child transmission, the CT values can provide insights into the risk of transmission from mother to infant. Typically, a higher CT value (indicating a lower viral load) would suggest a lower risk of MTCT for HIV-2. This distinction is important for tailoring prevention and treatment strategies specifically for HIV-2, considering its unique virological characteristics compared to HIV-1.

**Discussion:** This study marks a pioneering report on the possible transmission of HIV-2 from mother to child in Uganda, with 4 cases identified between May to November 2023. The HIV-2 positive samples exhibited high CT values, indicative of low viral load that is characteristic for HIV-2. Further investigations are ongoing to gather more details about the HIV-2 positive infants and to perform alternative tests to rule put non-specificity.

## Introduction

The Human Immunodeficiency Virus (HIV), an enveloped retrovirus, can be classified into two main types: HIV type (HIV-1) and HIV type 2 (HIV-2) [1]. In 1985, two years after the discovery of HIV-1, the existence of HIV-2 emerged due to suspicions arising from atypical serological patterns in Senegalese patients.[2] Though both viruses are transmitted through similar routes and can result in acquired immunodeficiency syndrome (AIDS), HIV-2 progresses much slower than HIV-1. Nevertheless, they significantly differ in aspects such as epidemiology, diagnosis, and treatment management. [3] HIV-2 accounts for 3 to 5% of the global HIV burden, with current estimates suggesting approximately 1-2 million infected individuals worldwide. [4] The highest prevalence rates of HIV-2 in the world were known be recorded in parts of populations in West African countries. [5, 6] Within Europe, France and Portugal have reported the highest prevalence rates of HIV-2 infection, with approximately 1000 and 2000 cases recorded in each country, respectively. [6]

The historical diagnosis of HIV-2 has faced challenges related to differentiating HIV antibodies and detecting plasma HIV-2 RNA levels. Clinicians used to rely on clinical signs to determine whether specific HIV-2 testing was necessary before the widespread availability of 4^th^ generation HIV-1/2 antigen-antibody tests. Earlier immunoassays could detect HIV-1 and HIV-2 antibodies but couldn’t distinguish between them. Confirmatory testing using western blots was often utilized but the overlap between HIV-1 and HIV-2 antibodies often led to inconclusive results or misdiagnosis as HIV-1. [1] In 2014, the U.S Centre for Disease Control (CDC) developed an algorithm for diagnosing HIV which included a supplemental test to differentiate between HIV1 and HIV-2.[7, 8]

For patients diagnosed with HIV through enzyme-linked immunosorbent assays and exhibit a low or undetectable HIV-1 viral plasma load without undergoing antiviral therapy (ART), it’s important to contemplate the potential existence of HIV-2 or other HIV variants. Additional testing using specific antibody-based confirmatory assays should be conducted in such cases. Moreover, patients undergoing ART with consistent CD4+ cell decline despite the absence of detectable HIV-1 plasma RNA should undergo re-evaluation for possible HIV-2 infection, co-infection of both HIV-1 and HIV-2, or the presence of other HIV variants. [9]

In Uganda, the shift from the CAP/CTM Qual HIV-1 assay, which has been in use since 2010, to the more discerning Cobas^®^ HIV-1/HIV-2 Qualitative nucleic acid test in 2023 marked a significant advancement in detecting and differentiating HIV types in infants. The Cobas^®^ HIV-1/HIV-2 Qualitative nucleic acid test for use on Cobas^®^ 5800/6800/8800 systems has the ability to differentiate between HIV-1 and HIV-2 to detect presence of HIV-2 in DBS, Serum, Plasma, and whole blood samples. This study aims to employ this advanced testing method to provide evidence of HIV-2 transmission from mother to child in Uganda, a critical step in enhancing the nation’s public health response to HIV.

## Methods

### Study design

This study conducted a retrospective analysis of existing routine laboratory data from early infant diagnosis (EID) programs in Uganda. This review of previously collected data enabled the identification and analysis of HIV-2 cases among infants born to HIV-positive mothers, providing insights into the virus’s prevalence and transmission patterns.

### Setting

Early infant diagnosis testing-methods within Uganda’s public health sector were established in 2010 using the CAP/CTM Qual HIV-1 Test and have been performed according to the Uganda consolidated guidelines for HIV prevention and treatment. The CAP/CTM Qual HIV-1 Test was replaced by cobas^®^ HIV-1/HIV-2 Qualitative nucleic acid test for use on cobas^®^ 5800/6800/8800 systems in May 2023. Testing is routinely performed using DBS specimens obtained from capillary heel-prick (approximately 70 μl per spot per test).

24,542 DBS samples were collected from infants below 18 months and born to HIV positive mothers. These samples were tested on cobas^®^ 6800 systems using the cobas^®^ HIV-1/HIV-2 Qualitative nucleic acid test during the period of May-November 2023 at Central Public Health Laboratories Uganda. The cobas^®^ HIV-1/HIV-2 Qualitative nucleic acid test is based on fully automated sample preparation (nucleic acid extraction and purification) followed by PCR amplification and detection. The cobas^®^6800 Systems consist of the sample supply module, the transfer module, the processing module, and the analytic module. Automated data management is performed by the cobas^®^ 6800 Systems sohware which assigns test results for all tests as non-reactive, reactive, or invalid. Results can be reviewed directly on the system screen, exported, or printed as a PDF report.

The results were retrieved from the shared server of Central Public Health Laboratories and entered into an excel document for analysis.

## Results

Of the 24,619 DBS tested between May and October 2023, 466(1.9%) were confirmed HIV positive. Of these, a smaller subset—four cases (0.9% of the positive samples)—were identified as HIV-2. These cases were distributed across four districts: Kampala (1), Kagadi (1), Namutumba (1), and Oyam (1) districts. Notably, the CT values for these HIV-2 positive samples were higher (39.7, 40.33, 40.35, 44.62), suggesting lower viral loads when compared to the HIV-1 samples, which typically showed CT values below 30. This pattern is consistent with the expected viral load profiles for HIV-2 infections and places these cases within the 95th percentile for CT values. Analysis of this data was carried out using Microsoft Excel.

| Sample ID | Result | Ct value | Date tested |
| --- | --- | --- | --- |
| 1 | HIV 2 Reactive | 44.62 | 07/23 |
| 2 | HIV 2 Reactive | 39.67 | 08/23 |
| 3 | HIV 2 Reactive | 40.33 | 11/23 |
| 4 | HIV 2 Reactive | 40.35 | 11/23 |

## Discussion

The findings presented in this study indicate the prevalence of HIV-2 among infants born to mothers infected with HIV in Uganda using the cobas^®^ HIV-1/HIV-2 Qualitative nucleic acid test on the cobas 6800 system. The study also shows that HIV-2 is less prevalent (0.9%) compared to HIV-1 among the tested infants, indicating that it is less common in this population. The utilization of the cobas^®^ HIV-1/HIV-2 Qualitative nucleic acid test underscores the critical importance of differentiating between the two viruses due to their differing risk of transmission, disease progression and treatment responses. Notably, the under-detection of HIV-2, particularly in regions where HIV-1 is predominant, could undermine control efforts if not adequately addressed.

The identified HIV-2 samples had high CT values in comparison with HIV-1 positive ones. This indicates the low viral loads of HIV-2 infection thus lower viremia. This is also consistent with the nature of HIV-2 infection, which is characterized by a slower progress to AIDS compared to HIV-1 infection. Furthermore, the distribution of identified HIV-2 cases across various regions of Uganda—namely Kampala, Kagadi, Namutumba, and Oyam—warrants further investigation to elucidate the factors influencing this spread.

The study’s results have significant implications for public health efforts in Uganda. The results reinforce the necessity for sustained vigilance in HIV surveillance but also highlight the need to evolve diagnostic capabilities to keep pace with the epidemiological nuances of HIV infections. Taking a cue from other nations that have encountered HIV-2, such as France and Portugal, Uganda could benefit from a more nuanced approach to managing HIV-2, incorporating targeted diagnostics and antiretroviral treatments tailored to its specific resistance patterns. This strategy would not only bolster the current preventive measures but also provide a more robust response to the diverse presentations of HIV within the population.

In light of our findings, ongoing investigations are imperative. We must continue to explore the specifics of HIV-2 positive infants and refine our testing methods to ensure specificity and sensitivity. As Uganda fortifies its fight against HIV, integrating lessons from international practices could enhance the efficacy of national initiatives, paving the way for a more informed and adaptive public health strategy.

## Conclusion

In conclusion, this study provides findings and evidence of presence of HIV-2 among infants born to HIV-infected mothers in Uganda, which advocates for continued vigilance in monitoring both HIV-1 and HIV-2 infections individually. The evidence suggests that while HIV-2 is less prevalent than HIV-1, its impact on public health is non-negligible. This, therefore, also urges policy makers and health practitioners to maintain a vigilant monitoring system that distinguishes between HIV-1 and HIV-2, ensuring that each virus is understood and managed effectively. A deeper understanding of both viruses will enhance the accuracy of diagnoses, refine treatment regimens, and support the ultimate goal of eradicating both forms of HIV in Uganda. By recognizing the distinct challenges posed by HIV-2, Uganda can develop a more comprehensive and responsive HIV control strategy that is informed by the nuances of both viral types.

## Ethical Considerations

A waiver of informed consent was received from Uganda National Health Laboratory Services Ethics and Research committee (UNHLSREC) to use data since no additional procedures or interventions were required from the participants, there was no risk identified.

## Data Availability

All data produced in the present study are available upon reasonable request to the authors

